# “It Is Better to Live Than to Die”: A Qualitative Study on Perceptions of Radiotherapy Among Patients with Cancer in Southern Ethiopia

**DOI:** 10.1101/2025.07.08.25331120

**Authors:** Dereje Geleta, Getahun Sintayehu, Netsanet Bogale, Betty Ferrell, Lesley Taylor, Andrew Tam

## Abstract

**Introduction:** Radiotherapy is one of the cornerstones of cancer treatment; however, access remains limited in many low-resource settings, particularly in low- and middle-income countries, such as Ethiopia. Recent efforts by the Ethiopian government have aimed to strengthen the oncologic infrastructure, including the building of new radiotherapy centers. Many non-structural barriers continue to exist, negatively impacting access. A better characterization of perceptions of radiotherapy is needed to guide future efforts in improving cancer outcomes.

**Objective:** This study aimed to explore perceptions of radiotherapy, including barriers to accessing radiotherapy, among patients with cancer receiving cancer treatment in southern Ethiopia.

**Method:** We conducted an explanatory qualitative study among patients with cancer receiving treatment at the Hawassa University Comprehensive Specialized Hospital Cancer Treatment Center (HUSCH-CTC) from March to May 2024. Data were collected through in-depth interviews guided by an open-ended interview guide and audio-recorded with participants’ consent. Transcripts from the interviews were then coded, and thematic analysis was performed.

**Result:** To our knowledge, this was the first qualitative study on patients’ with cancer perceptions of radiotherapy in southern Ethiopia. We found that participants generally held negative views about radiotherapy, which were shaped by multiple factors: concerns about side effects, uncertainty regarding treatment outcomes, and circulating misinformation and misconceptions within the community. A lack of or limited understanding of high-energy radiation (or X-ray) among fellow patients and the general public further reinforced these negative perceptions.

**Conclusion:** The findings from this study further support the need to empower patients via education and improve communications between healthcare providers and patients about radiotherapy and cancer treatments in general. Facilitation of education and dialogue is likely to increase awareness, dispel misconceptions, and thereby positively influence patient perceptions, informed consent, and adherence to radiotherapy to improve cancer outcomes.

## Introduction

Radiotherapy (also called radiation treatment or therapy) is a cancer treatment that uses high doses of radiation (such as X-ray) to target cancer cells and shrink tumors by damaging their DNA (1,2). The effect is not immediate but rather takes days or weeks to observe the treatment effects (1,3–5). Radiotherapy may be delivered externally (“external-beam radiation therapy”) or through internal radiation therapy (“brachytherapy”) (3,6).

Radiotherapy can be considered for almost all types of cancers with various treatment intentions, ranging from curative intent in many early-stage cancers or non-operative cases, adjuvant intent after surgery to reduce the risk of recurrence in the tumor bed or surrounding at-risk regions, and palliative intent to alleviate symptoms caused by cancer (7). The decision to employ radiotherapy, as well as its dose and fractionation, depends on many patient clinical factors such as the type and stage of cancer, performance status, and other medical comorbidities (2,3,7).

Accessibility and availability of radiotherapy are limited in many low- and middle-income countries (LMICs), where it is estimated that only ten percent of patients have access to radiotherapy (8). Among the 49 sub-Saharan African (SSA) countries, more than 20 countries currently lack any radiotherapy facilities (8–10). Furthermore, among the SSA countries with radiotherapy services, access to radiotherapy remains scarce, such as having only a single radiotherapy machine for geographic areas serving millions of people (11–13). Beyond the limited number of radiotherapy centers in LMICs, other multifaceted structural and social barriers were also identified by prior studies to be important factors negatively impacting access to radiotherapy, including: equipment shortage and high costs, insufficiently trained workforce, inadequate funding and overwhelmed health systems, logistic challenges, geographic challenges, and cultural factors., (14–17).

**Cancer rates in Ethiopia have climbed over the last ten years, reaching an estimated 53,560 new cases in 2019 for both men and women. The age-standardized incidence stood at roughly 104.5 per 100,000 population annually (18,19). Among the most frequently diagnosed cancers are leukemia, cervical, breast, colorectal, and stomach cancers. In women, breast cancer is the most common cancer diagnosis, accounting for about one-third of all female cancer cases, while colorectal cancer and non-Hodgkin’s lymphoma are particularly common in men (20)**.

In Ethiopia, radiotherapy is an essential component of cancer care in providing both curative and palliative care, but it is limited by the infrastructure. In addition, there are often very few oncologists for the entire population, leading to long waiting times and treatment delays. Furthermore, most patients with cancer present at advanced stages due to low awareness, lack of screening programs and poor referral systems. For example, in Ethiopia, the radiotherapy service had long been highly centralized in the capital city, primarily Tikur Anbessa Specialized Hospital (TASH) in Addis Ababa (21). In recent years, the government has actively expanded access to radiotherapy with the construction of radiotherapy centers in regional hospitals such as Gondar, Harar, Mekele, Jimma and Hawassa (12,22).

Prior studies from other low-income countries on patient perceptions of radiotherapy have revealed varying levels of fears, misconceptions, and awareness of radiotherapy. More importantly, these studies have also demonstrated that these factors could further negatively impact the uptake and adherence of radiotherapy. In Ethiopia, recent government efforts have aimed to expand the number of radiotherapy centers in the country. However, there is no comprehensive data about the perception of patients with cancer towards radiotherapy. This study aimed to explore the perceptions of radiotherapy among patients with cancer receiving treatment in southern Ethiopia.

## Methodology

### Study Setting

This study was conducted at Hawassa University Comprehensive Specialized Hospital-Cancer Treatment Center (HUSCH-CTC) in Hawassa from April to May 2024. Hawassa, located in southern Ethiopia, is the capital city of Sidama regional state. The city has various public and private health facilities with two primary hospitals-a general hospital and a comprehensive specialized hospital (a cancer treatment center). The HUSCH-CTC is the only public cancer treatment center in the city. Additionally, there are several private health facilities offering different levels of oncologic services.

HUSCH-CTC is the only regional comprehensive cancer treatment center, serving more than 25 million people in southern Ethiopia. Further details about HUCSH-CTC are available in a previous publication (23). Ethiopia, has only five radiotherapy centers for a population of more than one hundred million people, with 80% of whom reside in rural areas around the country. In recent years, the government has worked to strengthen the oncologic infrastructure by developing a National Cancer Control Plan, leading to progress in expanding the number of radiotherapy centers across the country. As part of this initiative, a radiotherapy unit was established at HUSCH-CTC and began offering radiotherapy in late 2024. At the time of data collection of this study, HUCSH-CTC radiotherapy service was in its initiation phase. For patients who were recommended to receive radiotherapy, patients were referred to the capital, Addis Ababa, for care.

### Study population and Sampling techniques

A maximum variation sampling technique was employed. We included participants with diverse backgrounds in terms of sex, age, cancer diagnosis, previous and current cancer-related treatments (including radiotherapy), and cancer stage. Patients under the age of 18 years or unable to provide consent were excluded. All participants were provided with an information sheet describing the background and objectives of the study. Consent to take part in the study and be audio-recorded were collected before proceeding with the in-depth interviews.

### Data collection

Data were collected via in-person in-depth interviews conducted by trained and experienced public health professionals. All interviews were conducted in the local language, Amharic. An interview guide with an open-ended question for each section, followed by a series of open- and closed-ended questions, was used to facilitate the interviews. The guide included the following sections: perceptions about radiotherapy, understanding about radiotherapy, communication about radiotherapy, barriers to radiotherapy, and patients’ recommendations about radiotherapy related communication. The interviews were conducted in a pre-arranged private room to ensure the participants’ comfort, privacy, and confidentiality. Participant demographics, including cancer diagnosis and stage, were recorded by reviewing the medical records of the participants. The interviews were audio recorded.

### Data analysis

All interview recordings were transcribed into English from the local language (Amharic). Codes were generated, and themes were developed. The data analysis was managed using ATLAS.ti,version 7.5 (ATLAS.ti Scientific Software Development GmbH, Berlin, Germany).

### Ethical consideration

Before the commencement of the study, ethical clearance was obtained from the Institutional Review Board (IRB) of Hawassa University (***Ref*.*No: IRB/216/16***). The study was carried out according to the principles stated in the Declaration of Helsinki, all applicable regulations, and according to established international scientific standards. Considering low levels of literacy in the study population, permission to use verbal consent was specifically provided by the ethics committees. Study teams read the study consent form to all prospective participants and obtained verbal permission to participate in the study. Each individual interviewed was anonymized via a specific code during transcription.

## Result

### Characteristics of study participants

A total of twenty participants were recruited and all consented to participate in the in-depth interviews. Demographics of these participants in terms of sex, age, history of radiotherapy, cancer diagnosis, treatment and stage of cancer are summarized in Table 1.

**Table 1.** Socio-demographic characteristics of participants.

| Variables | Response | Number (%) |
| --- | --- | --- |
| Sex | Female | 11 (55) |
|  | Male | 9 (45) |
| Age | < 50 years old | 12 (60) |
|  | ≥ 50 years old | 8 (40) |
| History of radiotherapy | Yes | 4 (20) |
|  | No | 16 (80) |
| Cancer diagnosis | Breast | 11 (55) |
|  | Lung | 1 (5) |
|  | Recto sigmoid | 1 (5) |
|  | Gastrointestinal | 4 (20) |
|  | Esophageal | 1 (5) |
|  | Lymphoma | 1 (5) |
|  | Ovarian | 1 (5) |
|  | Renal cell | 1 (5) |
| Treatment history | Attending Chemotherapy | 13 (65) |
|  | Undergone Surgery | 4 (20) |
|  | Not started | 2 (10) |
| Stage of cancer | Stage I | 1 (5) |
|  | Stage II | 3 (15) |
|  | Stage III | 6 (30) |
|  | Stage IV | 8 (40) |
|  | Not specified | 2 (10) |

**Table 2.** Illustrative quotes on negative and positive perceptions of the radiotherapy.

| *Quote* | *Data source* | *Participant* | *Theme* |
| --- | --- | --- | --- |
| *Because treating cancer cells with radiation also affects normal cells and other healthy parts of the body, I have significant apprehension about radiotherapy.* | *IDI* | *Woman, receiving chemotherapy treatment* | *Negative perception* |
| *I have a negative perception of radiotherapy because, during treatment, the radiation may damage adjacent healthy cells. Additionally, my friend nearly died as a result of radiotherapy.* | *IDI* | *Man, adult, renal cell carcinoma, treatment not yet started* | *Negative perception* |
| *It is difficult to hear the term "radiotherapy" because it has the potential to harm internal organs rather than just targeting the disease.* | *IDI* | *Woman, breast cancer, receiving chemotherapy treatment* | *Negative perception* |
| *I have heard negative things about radiotherapy, such as that once it starts, even healthy parts of the body can be affected. I have also heard that it could cause mental health disorders; therefore, if radiotherapy were recommended for me, I would be fearful of using it.* | *IDI* | *Woman, breast cancer, receiving chemotherapy treatment* | *Rumors, misconception* |
| *Based on the information I have heard, it is often said that those who undergo radiation do not achieve a good outcome in the end.* | *IDI* | *Woman, breast cancer, treated with surgery* | *Rumors, misconception* |
| *My positive perception is that radiotherapy may cure the disease because it burns the cancerous area and prevents it from growing and spreading.* | *IDI* | *Man, renal cell carcinoma, treatment not yet started* | *Positive perception* |
| *A patient with cancer told me that she was cured after undergoing radiotherapy treatment, so I do not have any negative considerations about radiotherapy.* | *IDI* | *Woman, ovarian cancer, Chemotherapy and surgery* | *Positive perception* |
| *Radiotherapy is one of the latest types of cancer treatment modalities that aims to eliminate all cancer cells from the body. It is often considered the last option for treating cancer.* | *IDI* | *Man, , Recto sigmoid cancer, surgery and chemotherapy* | *Positive perception; Misconception* |

**Table 3.** Illustrative quotes on perception before and after radiotherapy.

| *Quote* | *Data source* | *Participant* | *Theme* |
| --- | --- | --- | --- |
| *When radiotherapy was recommended to me, I was very scared. I thought it would be a difficult experience. I had the perception that it would cause burns, and until the moment I entered the machine, I was afraid it might cause severe pain. As to live it better than to die, I decided to take radiotherapy. it was a choice between life and death.* | *IDI* | *Woman, Breast cancer* | *Perception before radiotherapy* |
| *When I heard that[radiotherapy], I was very scared and started considering radiation(cherer). I thought the chances were fifty-fifty—it could go either way. I decided to accept whatever outcome came.* | *IDI* | *Woman, elder, breast cancer* | *Perception before radiotherapy* |
| *It was very difficult. I worried a lot and didn't think I would survive or even make it out of the therapy room. The fear was overwhelming. I decided to go through with the therapy because it was my only option.* | *IDI* | *Woman, Breast cancer* | *Perception before radiotherapy* |
| *Hair loss was caused by chemotherapy, which was very difficult for me. I experienced the side effects that the doctors had warned me about. Radiation, on the other hand, was much easier to handle. With God's help, I got through it* | *IDI* | *Woman, Breast cancer* | *Perception after radiotherapy* |
| *Chemotherapy's side effects were much harder to handle, such as hair loss, weakness, and body aches.* | *IDI* | *Woman, Breast cancer* | *Perception after radiotherapy* |
| *Chemotherapy was very, very difficult, but radiotherapy wasn't as bad.* |  |  |  |
| *It wasn't as bad as I expected. Chemotherapy, for example, causes more severe side effects, such as hair loss. I experienced more discomfort during chemotherapy than during radiotherapy. With chemotherapy, I lost my appetite and suffered more, but with radiotherapy, it wasn't as difficult.* | *IDI* | *Woman, Breast cancer* | *Perception after radiotherapy* |

### Key Themes

From this study, we developed 8 key themes: Perceptions of radiotherapy, concerns related to radiotherapy, comparison of radiotherapy to other treatments, communication about the details of radiotherapy, understanding of the role of radiotherapy and why radiotherapy was recommended, experience of radiotherapy, barriers and challenges, and family support related radiotherapy.

### Perceptions about Radiotherapy

The participants expressed negative perceptions about radiotherapy (locally known as *cherar hikimina*). Although they expected radiotherapy to lead to better outcomes for their cancers (perceived positive outcome), many also voiced concerns that patients treated with radiotherapy might eventually develop other diseases (perceived threat). One of the most emphasized fears was that radiotherapy might damage normal cells, leading to harm to various body parts. Participants noted that radiotherapy could penetrate the body and thereby could adversely affect internal organs. Furthermore, it was perceived that, given the high penetration power, radiotherapy was difficult to control (perceived threat) and could lead to unintended damage to healthy organs (perceived severity). This negative view of radiotherapy was largely attributed to the radiation (locally known as *cherer*), which carries a negative connotation in the community, used during treatment..

The participants attributed the negative information circulating within the community to have led many people to harbor fearful and negative perceptions of radiotherapy. Beliefs persist that radiotherapy eventually results in disabilities, such as loss of balance, loss of sight (blindness), and mental problems (memory loss).

Moreover, it was highlighted that radiotherapy was recommended for patients with advanced stages of cancer, when other treatment methods might no longer be helpful. Therefore, for many, radiotherapy was viewed as a temporizing measure for these stages. Consequently, radiotherapy was often seen as an indicator of the patient’s level of disease severity. In other words, radiotherapy was seen as a “last resort” when there were no other treatments available.

Radiotherapy was also perceived by some as the latest and most effective cancer treatment method, leading to better outcomes (perceived benefit) despite its side effects (perceived barrier). It was further noted that if radiotherapy is recommended at an early stage of the disease, it results in a better prognosis.

### Concerns Related to Radiotherapy

A lack of clear understanding and adequate information about radiotherapy was pointed out during the discussions. One common misconception is that radiotherapy, if administered during the early stages of the disease, could damage other body organs. Additionally, there are concerns that radiation used to treat the existing cancer may cause another cancer to develop. Other concerns expressed by these participants about radiotherapy include its adverse side effects, the occurrence of scars, and negative rumors and misconceptions regarding the treatment.

### Radiotherapy Compared to Other Common Cancer Treatments

Diverse opinions emerged regarding radiotherapy when compared with other cancer treatments. Participants primarily compared side effects, effectiveness, administration mode, and treatment accessibility among the different cancer treatments. Some stated that radiotherapy was more effective than other treatments, and thereby justifiable for its limited accessibility and high expense. Some compared it with chemotherapy alone, noting that while the “chemicals” from chemotherapy stay in the body, radiotherapy is administered quicker and more effective in killing cancer cells. However, a large number of participants mentioned that chemotherapy and surgery were more effective than radiotherapy. They also emphasized that they considered radiotherapy to have more side effects when compared with chemotherapy and surgery. Although participants experienced various side effects during chemotherapy, they perceived the side effects of radiotherapy as more severe. For instance, participants highlighted chemotherapy is administered intravenously, its side effects are not visibly apparent, whereas radiotherapy often leaves a visible scar and may damage normal or healthy organs.

The perceived side effects of radiotherapy were thought to be more severe than the actual side effects experienced from other treatments, such as surgery and chemotherapy. Conversely, it was noted that surgery involves pain and wounds, and chemotherapy is associated with significant side effects such as weight loss, loss of appetite, nausea, vomiting, and stress, all of which are perceived to be less severe compared to those side effects associated radiotherapy.

### Communication about Radiotherapy

Conversations about radiotherapy were uncommon. The majority of participants emphasized that they rarely discussed radiotherapy with anyone, stating that such communication was unnecessary unless the treatment was recommended for them—a sentiment attributed to the fears surrounding radiotherapy. Moreover, the overall negative perception of cancer within the community led to barriers to open dialogues about radiotherapy and other cancer treatment methods. It was noted that it was common for individuals who were diagnosed with cancer to not disclose their condition to their community, and even healthy individuals felt uncomfortable discussing radiotherapy.

Participants conveyed that discussions about radiotherapy mainly occurred among patients with cancer themselves. Participants highlighted that they engaged in discussions with others who had undergone radiotherapy. Such discussions, most often centered around side effects and treatment effectiveness, were noted to have a positive influence on those without prior experience with radiotherapy. In contrast, many commented that there were limited communications about radiotherapy between patients and health care providers. Only a few participants acknowledged having discussions with health care providers regarding the treatment, and these discussions were often viewed as inadequate.

There were, however, demands for more detailed discussions about radiotherapy with health care providers. Participants expressed a preference for receiving information about radiotherapy and other treatments directly from doctors, as information obtained from other patients or external sources was considered less accurate and more subjective.

### Understanding Why and When Radiotherapy Is Recommended

Both participants who had been treated with radiotherapy and those who have not yet been recommended for radiotherapy held unclear understandings of why radiotherapy was recommended. Many participants expressed discomfort and a sense of hopelessness when the radiotherapy was recommended. This was attributed to some widely circulated information that radiotherapy was seen as a last-resort option recommended only in the latest stages of disease. A few participants mentioned that radiotherapy was recommended to control the spread of disease, to kill cancer cells, and to supplement surgery and chemotherapy to optimize treatment outcomes, although this perspective was held by only a small proportion of participants.

### Experience Before and After Radiotherapy

Prior to undergoing radiotherapy, many participants expressed that they experienced fear and concerns regarding the treatment. More notably, the recommendation of radiotherapy induced a profound sense of hopelessness. Many had to face the difficult decision whether to accept or decline the treatment - it was perceived as a choice between life and death. However, nearly all participants emphasized that they chose to proceed with radiotherapy because they perceived radiotherapy as their only option, doing so with the hope that the treatment would cure the disease (perceived benefit). Some, however, questioned why the disease was not cured despite completing all recommended radiotherapy sessions.

After completing radiotherapy, most participants reported positive feelings. Contrary to their initial concerns, they did not experience the major side effects that they had feared initially. Instead, the side effects were evaluated as minor when compared to those experienced from other treatments, such as chemotherapy. Common side effects attributed to radiotherapy included nausea and vomiting (or food intolerance), patches of darkened skin around the treatment site, weight loss, and mild fatigue. Importantly, these side effects were seen as temporary and to not have long-lasting impacts.

Furthermore, participants acknowledged that, while radiotherapy did not cure the disease, it significantly improved their overall outcome. Overall, those who received radiotherapy reported a positive change in their perception of the treatment after having completed radiotherapy.

### Barriers and Challenges to Radiotherapy

Several factors were identified as potential barriers and challenges to receive radiotherapy. Among the participants, there was a strong perception that accessibility of radiotherapy care was a major challenge. Some participants noted that they were not aware of the locations of radiotherapy centers throughout the country. Participants also expressed that the cost of transportation, especially when the radiotherapy centers were not locally available, added to the perceived challenges of obtaining radiotherapy. Additionally, participants also mentioned that the expense of lodging during the treatment course further burdened patients, particularly as they often had to be accompanied by someone.

> *“I had to stay for ten days before the treatment began, and the treatment itself lasted for four consecutive weeks. During that time, I spent a significant amount of money. It was a major challenge for me, and I made many sacrifices—even selling my assets. I lost everything I had, hoping to be cured of this disease once and for all*.*” (woman, adult, attended radiotherapy)*
>
> *“I spent more than 72,000 ETB (approximately 1,385 USD) for one course of treatment, not including additional costs such as transportation and food. There were also delays—the service was sometimes not provided on the scheduled day*.*”(Woman, adult, attended radiotherapy)*.
>
> *“The cost of cancer treatment can be a significant financial burden*.*” (woman, Adult, attended radiotherapy*

Long wait times to receive radiotherapy meant additional travel and lodging expenses, and treatment schedules were often disrupted. Participants experienced repeated postponements and multiple trips to radiotherapy centers, which exacerbated the overall expense. This issue was particularly acute for centers located in Addis Ababa, the capital, where the cost of living is high, and for centers such as those in Jimma and Haramaya, which are far from the city center. Participants who underwent radiotherapy emphasized that the financial challenge during the course of treatment was compounded by social and psychological challenges, as both patients and their accompanying family members sometimes missed or even lost their jobs due to the prolonged treatment period.

> *The main challenge was not being able to receive treatment on my scheduled appointment dates. As patients, we’re often moved from one place to another, and there are many procedures to go through. I now have follow-up appointments every three months (women, adult, attended radiotherapy)*
>
> *It was very difficult. There were many patients at the radiotherapy centers, and people like us, who come from rural areas, face numerous challenges (women, adult, attended radiotherapy)*
>
> *My husband left his job to support me, which had a financial impact on our family. During those ten days, we also had children at home who needed care (women, adult, attended radiotherapy)*

### Family Support

Family support was acknowledged as a critical factor throughout the course of any cancer treatment. However, the prolonged duration of cancer treatment posed a challenge in maintaining the same level of family support. It was noted that the economic and employment status among their family members would often impact the level of support available. Participants noted that they received support from their children, relatives, and spouses, but recognized that such support became more challenging during radiotherapy due to the significant financial costs associated with it.

> *“My children supported me. They covered all my expenses*.*(Women, adult, attended radiotherapy)*
>
> *“I had support when I first started treatment, but now I don’t. I am facing challenges. My husband is unemployed, and I am covering the costs with my own work. If radiotherapy is recommended, it will be a major challenge for me*.*” (Women, adult, attending chemotherapy)*
>
> *“I have strong family support, but if I’m forced to travel far for radiotherapy, it will be challenging and I’ll become a burden to my relatives” (Man, elder, not started attending treatment)*

## Discussion

In this qualitative study using in-depth interviews, we explored the perceptions among patients with patients towards radiotherapy. To our best knowledge, this was the first study that aimed to assess these perceptions in Ethiopia, and one of the few in Africa. Overall, we found four main key themes: (1) negative perceptions about radiotherapy; (2) mixed beliefs about its efficacy compared with other treatments; (3) concern over side effects, and (4) association with advanced disease severity.

One of the interesting findings that was not reported in prior studies, was that many participants believed undergoing radiotherapy to be an indicator of advanced cancer and thus viewed by the community as a treatment of last resort. These perceptions contributed to the feelings of hopelessness, stress, and anxiety among the patients at the time when radiotherapy was recommended. As a result, for many patients, the decision to receive radiotherapy was considered a choice between “to live or to die”.

Across the interviews, patients reported an absence or minimal patient-health care provider communications regarding why radiotherapy was recommended and how it works. This lack of patient-provider communication created a significant gap about the treatment. **Most participants described themselves as passive recipients of care, relying on information from other patients, family members, and the general population information that is not always correct. Such individually driven information can expose patients to different concerns**. We believe that improved understanding of the perceptions among patients with cancer toward radiotherapy is essential in making informed decisions for radiotherapy and improving its uptake and adherence. In settings where many patients reside in rural and low literacy areas, understanding the perceptions of patients and the general public is important to improve the uptake and adherence to treatment.

Rumors and misconceptions about radiotherapy were consistently mentioned as key drivers of patient anxiety. Participants expressed fear that radiation would burn them and destroy internal organs and other healthy parts of the body. Many assumed the presence of radiation alone would cause harm. This narrative is reflected in deep-seated beliefs within both patients with cancer and the general population.

Limited access to radiotherapy centers, extended waiting times, and financial burdens associated with travel and lodging were emphasized as barriers. These logistical challenges reinforced negative perceptions toward radiotherapy and discouraged patients from pursuing and adhering to radiotherapy treatment.

Our findings are aligned with previous studies conducted in Ethiopia and other parts of Africa. A study from Tanzania indicated that about two-thirds of the public held a negative perception of radiotherapy (24,25). Fears of side effects are highly emphasized, leading patients to develop pronounced anxiety about radiotherapy. Studies conducted in Ethiopia have shown that cancer patients experience significant fear and anxiety about radiotherapy, shaped by misconceptions that it “burns” the body, irreparably damages internal organs, or invariably leads to death or poor outcome(26–28).

Moreover, several major concerns influence perceptions of radiotherapy: lack of trust in treatment outcomes; limited understanding of when and why radiotherapy is recommended; passive patient participation in decisions about treatment modalities; mixed opinions on radiotherapy’s effectiveness relative to other treatments; and low public awareness of cancer generally. Likewise, other studies have emphasized that limited awareness and education, cultural beliefs, associations with poor outcomes, psychological distress, and barriers to access are among the factors shaping patients’ with cancers decisions and willingness to consent to radiotherapy (12,14,24,28–30).

Our study also identified practical barriers: the inaccessibility of radiotherapy centers, long waiting times, and costs associated with travel and lodging were repeatedly cited as major obstacles. Consequently, these barriers foster negative attitudes and reluctance to consent to radiotherapy (12,14,24,27,28,31). Communication gaps between patients and healthcare providers emerged as another critical issue. Limited or absent discussions about cancer treatments leave patients without clear information, forcing them to rely on rumors and misunderstandings that negatively impact their perceptions and treatment decisions. While time constraints among caregivers may contribute to this lack of communication, participants consistently expressed a desire for accurate information from their providers.

By focusing on cancer patients’ perceptions of radiotherapy, this study highlights pervasive negativity and misunderstanding about the treatment. These attitudes stem from a combination of insufficient awareness regarding radiotherapy’s purpose and outcomes, accessibility issues, treatment costs, and poor patient–provider communication. Our findings underscore the practical importance of enhancing patient education about radiotherapy and strengthening communication skills among cancer care providers to bridge informational gaps.

A limitation of this study includes a small number of participants, a non-random sample drawn from a single cancer treatment center, which may limit generalizability. Additionally, Hawassa University Comprehensive Specialized Hospital’s Cancer treatment center radiotherapy service was in its initiation phase during data collection. Therefore, findings should be interpreted cautiously, especially when comparing to the current state of the facility.

We recommend that future research build on these insights by employing larger, more representative samples and participatory methods such as implementation research and human-centered design to develop localized, context-specific solutions. Such approaches could foster positive perceptions, improve understanding of radiotherapy, and enhance patient–provider communication about radiotherapy and others cancer treatments.

In conclusion, our study underscores that negative perceptions toward radiotherapy, driven by multifaceted factors, exist among cancer patients in Ethiopia. These insights deepen our understanding of how gaps in patient education and provider communication shape patients’ willingness to consent to and adhere to radiotherapy treatment.

## Data Availability

All data and related metadata underlying the findings reported in a submitted manuscript are already provided as part of the submitted article.

## Acknowledgment

The authors express their sincere gratitude to the administration and staff of the HUSCSH Cancer Treatment Center for their invaluable support and facilitation during the data collection period. We also extend our heartfelt appreciation to the interview facilitators for their willingness, time, and significant contributions to this study.

## Authors contribution

### Conception and design

Andrew Tam, Dereje Geleta, Netsanet Bogale, Getahun Sintayehu, Betty Ferrell, Lesley Taylor

### Financial Support

City of Hope National Medical Center through philanthropic donations. The funders had no role in study design, data collection and analysis, decision to publish, or preparation of the manuscript.

## Declaration of conflicting interests

The author(s) declared no potential conflicts of interest with respect to the research, authorship, and/or publication of this article.

### Administrative support

Netsanet Bogale, Lesley Taylor, Dereje Geleta, Andrew Tam

### Collection and assembly of data

Dereje Geleta, Getahun Sintayehu

### Data analysis and Interpretation

Dereje Geleta, Getahun Sintayehu

### Manuscript writing

All authors

### Final submission

All authors

### Accountable for all aspects of the work

All authors

